# Posterior Parietal Cortex Modulates Perceptual Decisions Depending On Psychotic Phenotype

**DOI:** 10.1101/2024.11.08.24316920

**Authors:** Francesco Scaramozzino, Ryan McKay, Nicholas Furl

**Author notes:** Corresponding author: Francesco Scaramozzino; Egham Hill, Egham TW20 0EQ, UK.

## Abstract

**BACKGROUND:** Reduced data-gathering and altered sensory precision are associated with psychotic phenotypes in tasks engaging the posterior parietal cortex (PPC). We investigated whether PPC excitability - modulated via 1 Hz repetitive transcranial magnetic stimulation (TMS) - differentially affects these behavioural patterns in high vs. low psychotic phenotypes. Based on prior work, we hypothesised that delusional and hallucinatory traits would moderate TMS effects on sensory precision (proxied by drift rates), while hallucinatory traits would additionally moderate effects on decision thresholds.

**METHODS:** We compared performance in both the random dot motion task (RDM) and the beads task in two groups of participants (total, *N* = 68) undergoing TMS or sham-TMS over the right PPC. Hierarchical drift-diffusion models estimated drift rates (sensory precision proxies) and decision thresholds. We evaluated differences between TMS and sham-TMS groups and tested for interactions of these TMS groups with delusional and hallucinatory phenotypes.

**RESULTS:** In RDM, TMS increased decision thresholds compared to sham-TMS in the low psychotic phenotype group. This effect was not present in participants with higher psychotic phenotypes. Drift rates, in contrast, were lowered in participants with higher delusional phenotype. No significant effect was found on beads task performance.

**CONCLUSIONS:** Our findings suggest a causal role of PPC in decisions to end data-gathering during perceptual inference. The absence of this effect in the psychotic phenotype yields new hypotheses on the role of PPC excitability in neural mechanisms underlying decision-making patterns in the psychotic phenotype.

## 1. Introduction

Theoretical and empirical work put forward the idea that our representations of reality emerge from neural inferences on “raw data” coming from the sensorium^1–3^. Alterations of these inferential mechanisms could produce unlikely inferences about reality, namely hallucinations and delusions^4,5^. Research on subclinical psychotic traits in the general and at-risk populations shows that the psychotic phenotype is present in a continuum^6–8^. Behavioural and computational patterns associated with altered inference were shown to vary along this continuum^9–13^. However, the neural mechanisms underlying this variability in behaviour and phenomenology are still unknown. The predictive coding account of psychosis (PCA) provides a theoretical framework linking phenomenology, computational and neural mechanisms^4,5^.

Based on the PCA, alterations of precision encoding should impact the agent’s representation of reality. Precision (i.e. inverse of variance) is a quantification of uncertainty and would be implemented in neurons by their synaptic gain, which is the probability of a presynaptic event causing a postsynaptic one^14^. Undue variations in precision (and so in synaptic gain) of prior or new evidence signalling could produce drastic changes in posterior estimations resulting in psychotic experiences. Using different modelling approaches in perceptual and probabilistic decision-making, numerous studies have shown that parameters that can approximate precision encoding vary with psychotic and psychotic-like symptoms^9,11,13,15–19^. According to the PCA, this variability in precision encoding should be coupled with variability in synaptic gain of neural circuits relevant to decision-making.

In paradigms requiring sampling of evidence, synaptic gain of posterior parietal cortex (PPC) superficial pyramidal neurons might be involved in alterations of precision encoding and quantity of accumulated evidence. Firing rates in the lateral intraparietal area (LIP) of monkeys performing the random dot motion task (RDM) increase with the quantity and quality of accumulated evidence until they reach a canonical threshold value just before the decision^20–22^. In humans, an analogous pattern has been found using electroencephalography (EEG): a positive centroparietal event-related potential known as centroparietal positivity whose amplitude increases with the quality and quantity of accumulated information in data-gathering tasks^23,24^. The centroparietal positivity seems to occur irrespective of sensory modality or motor execution and has been replicated in various paradigms including RDM^24–27^. Interestingly, the rate of ramping neural activity in the parietal cortex, measured via magnetoencephalography during the RDM, correlated with the synaptic gain of parietal cortex neurons, as estimated by dynamic causal modelling^28^. This suggests that neurons in the parietal cortex might encode the precision of accumulated evidence and signal when to stop gathering data.

In a previous study using RDM in combination with drift-diffusion models (DDMs), we associated both delusional and hallucinatory phenotypes with increased precision of sensory evidence and the hallucinatory phenotype only with lower decision thresholds^29^. We proxied precision of sensory evidence with the drift rate parameter in DDMs fitted on participants’ performance of the RDM^30^. The variability in precision encoding of sensory evidence associated with the psychotic phenotype that we found in RDM could reflect differences in the synaptic gain of PPC superficial pyramidal cells.

In the present work, we investigated whether changes in the excitability of PPC impact DDM parameters from RDM performance differently in individuals showing high vs. low psychotic phenotypes. We reduced excitability in the PPC surrounding the right intraparietal sulcus (rIPS; thought to be the human homologue of LIP in non-human primates^20^) through transcranial magnetic stimulation (TMS). Given the difference in sensory precision encoding between low and high psychotic phenotypes we previously found in RDM^29^, we expected both delusional and hallucinatory phenotypes to moderate any effect of TMS on sensory precision (proxied by drift rate). Given our previous results on hallucinatory phenotype affecting decision thresholds, we also expected hallucinatory phenotype to moderate any TMS effect on decision thresholds. Given the uncertainty associated with the effect of 1 Hz repetitive TMS on a cortical network level^31^, and consequently on behavioural performance, in both cases, we did not have any hypotheses about the direction of the moderation.

PPC activity has also been correlated with evidence accumulation and evaluation in a classic probabilistic reasoning task popular in psychosis research: the beads task (BT). Furl & Averbeck (2011) found higher BOLD responses in the rIPS in participants who decided to collect more evidence when the task uncertainty increased^32^. This link between PPC and evaluation of evidence uncertainty is supported by O’Reilly et al. (2013) who observed increased BOLD responses in IPS associated with higher trial uncertainty in a saccadic planning task^33^. This evidence suggests that the rIPS might be involved in the encoding of uncertainty associated with collected evidence in cognitive inference and in the decision to stop data-gathering at the cognitive or doxastic level, as opposed to the perceptual level tested using RDM. Given this evidence linking PPC and BT performance, we also investigated the effect of TMS on the quantity of collected evidence in BT and any interaction with psychotic phenotypes.

## 2. Methods and Materials

### Participants

Participants were recruited through the Sona Systems platform at Royal Holloway, University of London and were rewarded for participation with a £15 *Amazon* voucher or 3 university credits. Our study was approved by the ethics committee of Royal Holloway, University of London. After a complete description of the study, the participants could ask additional questions via email and choose to take part in the study with their informed consent. For reasons of TMS safety, we excluded participants who confirmed at least one of the following: a history of neurological or psychiatric conditions; having suffered from epilepsy, febrile convulsions in infancy or had recurrent fainting spells; having relatives suffering from epilepsy; having undergone neurological surgery; a heart pacemaker, cochlear implant, medication pump, surgical clips fitted in their body; taking any unprescribed or prescribed medication; undergoing anti-malarial treatments. We recruited only right-handed participants to control for hemispheric lateralisation.

Participants were invited to complete the first online part of the experiment through a link sent via email. The online part of the study was designed on the Gorilla platform and included: information about the study, the screening questionnaire, demographic questions (age and gender), the Peters et al. Delusion Inventory (PDI)^34^ and the Cardiff Anomalous Perception Scale (CAPS)^35^. Participants who matched at least one of our exclusion criteria on the screening questionnaire could not continue with the online study. After the completion of the online part, participants were invited to our facilities at Royal Holloway, University of London, where they underwent the in-person part of the study. In the laboratory, participants underwent our TMS protocol and just after the TMS session, performed each of the three behavioural tasks in a randomised order on a computer in the same room. After the screening, the number of participants who took part in the experiment was 70. However, because the online data of two participants were missing due to incompletion, the final total number of participants in our analysis is 68.

### Psychotic phenotype questionnaires

We used the 21-item PDI^34^ for evaluating the delusional phenotype and the CAPS^35^ for the hallucinatory phenotype. For both scales, we used the total score (sum of subscales). Using the median as the cut-off value, participants were categorised into two groups for PDI (high or low PDI; median= 60) and CAPS (High or low CAPS, median= 53).

### Behavioural tasks

We designed the RDM, BT and a simple reaction time task (SRT) using *Psychopy3* (v2021.2.3) (Peirce et al., 2019). The tasks were delivered in one *Psychopy3* experiment randomising the order of the tasks. All participants performed the tasks on the same computer and screen. For every task, each trial began with a white fixation cross (0.2×0.2 degrees of visual angle).

#### RDM

On a grey background, we presented 500 white dots within a 5-pixel radius. The dots were moving at a speed of 3 pixels/frame in a circular aperture with a 600-pixel diameter. Dots had 50 frame lifetimes, after which they were redrawn within the aperture. In all trials, the majority of the dots were moving randomly (noise dots) while a subset of dots moved coherently to the left or the right depending on the trial (signal dots). The noise dots were set to move in a random direction at each frame. The signal dots moved coherently for their entire lifetime. Participants performed four practice trials where the proportion of signal dots was 25% or 50%. Participants then completed 40 trials of two different coherence conditions in random order: 20 trials of a high coherence precision condition (HP) with 15% of signal dots; and 20 trials of a low coherence precision condition (LP) with 5% of signal dots. On each trial, the coherent motion of the dots was randomly assigned to be left or right. Participants ended the trial by indicating whether the signal dots were moving to the left or the right. To control the effects of the TMS on motor execution, participants were instructed to use only their left hand to respond and to place their index and middle fingers respectively on the “A” and “S” buttons for left and right. Participants were asked to answer as quickly and accurately as possible. After each trial, participants were asked to report confidence in their response with a slider on a 0-10 continuous scale.

#### Simple reaction time task (SRT)

We designed the SRT to control for any effects of TMS on motor execution and side of stimulus presentation (contralateral or ipsilateral to the TMS stimulation site). On each trial, a green circle of 250 pixels radius was presented randomly on the left or right of a grey background. As for RDM, to examine motor execution of the right hemisphere (targeted by TMS), participants were instructed to use only their left hand and to press the “A” button when the circle appeared on the left and to press the “S” button when the circle appeared on the right. Participants completed 30 trials where the circle could appear with a 50% probability to the left or to the right.

#### BT

We designed a draws-to-decision BT with an 80/20 colour ratio and a maximum of 10 beads drawable per trial. With the support of on-screen images, participants were encouraged to imagine two urns: a blue urn with 80 blue beads and 20 green beads, and a green urn with 80 green beads and 20 blue beads. At each bead sequence, they had to draw beads from a hidden urn before and guess whether it was the blue or the green urn. At each draw of the sequence, they were instructed to press the “1” button to guess the blue urn, “2” for the green urn and “3” to draw another bead. After the tenth bead was drawn, participants needed to choose one urn to continue to the next sequence or the end of the study. Before the experimental sequences, participants completed four practice sequences with an 80/20 colour ratio. After practice, participants completed 20 sequences of which 10 were blue urn sequences and 10 were green urn sequences presented in random order. At each draw, the colour of the bead shown on the screen was determined using a function that randomly picked with replacement one colour (“green” or “blue”) from a list of 100 strings of colour names where, depending on the trial, “green” and “blue” strings were present according to the 80/20 ratio. After each trial, participants were asked to report confidence in their response with a slider on a 0-10 continuous scale.

### TMS protocol

For each participant, we first localised P4 using the 10-20 system which was shown to reliably correspond to the cortical area around the rIPS^36^. Using a *Magstim Rapid²* machine, we established the individual active motor threshold (AMT) by stimulating the left primary motor cortex. The stimulation for AMT started at 50% of stimulation intensity, we then increased the intensity until a twitch in the participant’s right-hand fingers was induced. Participants were randomly assigned to a session of either 1 Hz TMS or the sham-TMS over P4. The intensity was set at 90% of the AMT intensity. During the 1 Hz TMS, magnetic pulses were delivered at 1 Hz frequency for 15 minutes. For the sham-TMS session, we reversed the TMS coil and delivered pulses at 1 Hz frequency so that the noise and the touch experience were preserved without the effect of the magnetic stimulation.

### Statistical analysis and computational modelling

#### Demographic group comparisons

We used the *Tableone* package in Python (Pollard et al., 2018) to test for the effects of TMS conditions, PDI and CAPS groups on age (independent T-test), gender (Chi-square), PDI and CAPS scores (Kruskall-Wallis test).

#### HDDM

We used the *HDDM 0.8.0* toolbox (http://ski.clps.brown.edu/hddm_docs/index.html)^37^ in an *Anaconda Python 3.6* environment to fit hierarchical Bayesian DDMs to RDM and SRT data. The DDM models binary decisions as stochastic sampling processes that take place in a continuous time frame. The DDM describes the increment of a decision variable *X* on a space of possible values of *X* over the time *t*. The inputs of the DDM are then the *RTs* and the value of *X* that can be either the accuracy (error or correct responses) or the binary value of a stimulus property (e.g., left or right). The increment 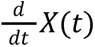 is determined by a stochastic process where values are sampled from a Gaussian distribution *N*(*v*, σ^2^). The accumulation process starts at an origin point where *t*=0 and *X* is equal to a given value *z.* For unbiased decisions, *z*=0; while in biased decisions z can assume positive or negative values if respectively biased towards decision *A* or *-A*. Depending on the precision of *N* (i.e., on 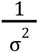), *X* would reach one of the two value options more or less quickly. The drift rate *v* quantifies this speed or rate at which *X* increases towards *A* or *-A*. The accumulation process ends for one option or the other when the value of *X* at time *t* is equal either to *A* or *-A.* The distance between *A* and *-A* in the *X/t* space is the decision threshold *a,* which indicates the amount of evidence needed for the accumulation process to reach a decision.

The estimation of DDM parameters in HDDM is embedded in a hierarchical Bayesian scheme where posterior estimates of parameters are obtained by using population-level prior parameter distributions (see supplementary material in Wiecki et al., 2003, for values of informative priors) and estimates of parameter distributions obtained by the Markov Chain Monte Carlo (MCMC) sampling method. We fitted RDM data to six different HDDMs where the parameters were free to vary for conditions/groups as illustrated in *Table 1*.

**Table 1.** HDDM specifications. Conditions/groups according to which drift rate, decision threshold and non-decision time parameters were free to vary. HC=High coherence LC=Low coherence

| Models | Condition/ Groups |
| --- | --- |
| Null-model | None |
| HDDM-C | HC vs. LC |
| HDDM-TMS | TMS vs. Sham |
| HDDM-TMS-C | TMS vs. Sham \| HC vs. LC |
| HDDM-TMS-PDI | TMS vs. Sham \| Low-PDI vs. High-PDI |
| HDDM-TMS-CAPS | TMS vs. Sham \| Low-CAPS vs. High-CAPS |
| HDDM-TMS-PDI-C | TMS vs. Sham \| Low-PDI vs. High-PDI \| HC vs. LC |
| HDDM-TMS-CAPS-C | TMS vs. Sham \| Low-CAPS vs. High-CAPS \| HC vs. LC |

We fitted SRT data to two HDDMs: HDDM-SRT-TMS, where parameters were free to vary across the TMS versus sham sessions; and HDDM-SRT-Null where one parameter value was fitted for both TMS conditions. For RDM, *X* was coded as accuracy values. *X* was coded as left or right choice for SRT, as SRT generally gives near-perfectly accurate performance and as we wanted to control evidence accumulation toward stimuli presented contralateral or not to TMS stimulation site rather than towards stimuli representing accurate or inaccurate response.

We ran 20,000 samples of MCMC iterations for each HDDM, discarding the first 2,000 samples as burn-in and using a thinning factor of 5 and a 10% outliers setting. Convergence of MCMC iterations was assessed by visual inspection and by computing the Gelman-Rubin statistic and verifying that values ranged between 0.9 and 1.1. We used the HDDM package to estimate and compare at the condition/group level the following DDM parameters: drift rate (*v*), decision threshold (*a*) and non-decision time (*t*). Since no prior bias has been induced in the task, we assumed *z*=0.

The best-fitting model was evaluated by comparing the deviance information criterion (DIC) obtained for each model. DIC is a Bayesian measure of fit which trades off the fit of the model and model complexity. Lower values of DIC suggest a better fit^38^. We considered significant a difference (Δ) of 10 DIC points^38^. By penalising model complexity, this DIC-based model selection decreases the baseline reliability of models testing multiple hypotheses (i.e., models with multiple comparisons or predictors), addressing the problem of multiple comparisons in a Bayesian context^39^. Because HDDM computes the posterior probability distributions of parameters for each condition or group, comparing them and determining the likelihood of a parameter being higher or lower in one condition or group than another is straightforward. We considered a difference between phenotype groups or conditions as statistically significant when the probability (*P)* of a parameter value being higher or lower in one condition/group compared to the other was >0.95^37^. For significance to look familiar to a frequentist eye, we reported significant results as the *(1-P)* probability being <0.05.

#### Linear mixed models

We used the *Statsmodels 12.2 toolbox* (https://www.statsmodels.org/dev/index.html) and evaluated the effect of TMS on DTD in BT using a linear mixed model where the TMS condition was set as the predictor and random slopes and intercepts were estimated for each participant.

We then employed hierarchical Bayesian regression analyses utilising the *PyMC3 3.11.2* software package in an *Anaconda Python 3.8* environment to examine the effects of TMS, PDI and CAPS on participants’ trial-level DTD. We fit four separate generalised mixed models with a Poisson distribution as a link function with setting predictors specified by *Table 2* (see *Supplementary Materials* for model specifications). We assigned to the intercepts’ prior distributions a mean centred on the median value of DTD and a standard deviation of 5. For the *β* coefficients, we employed relatively weak priors with a centred distribution around 0 and a standard deviation of 10. To account for individual variability in intercept and coefficient, we made these estimates vary for each participant.

**Table 2.** Bayesian regression models draws-to-decision. Bayesian regression linear models (LMs) predicting draws-to-decision in the beads task. x= interaction

| Regression Model | Predictor |
| --- | --- |
| Intercept-model | None |
| LM-TMS | TMS vs. Sham |
| LM-TMS×PDI | TMS, PDI, TMS × PDI |
| LM-TMS×CAPS | TMS, CAPS, TMS × CAPS |

We assessed the model fit by examining the Widely Applicable Information Criterion (WAIC) computed for each model^40^. The WAIC provides a measure of a model’s goodness-of-fit by quantifying its ability to predict new data points while also penalising for model complexity. Lower WAIC values indicate a better fit. By following the rule of thumb used for the DIC, a difference (Δ) of 10 WAIC units was deemed significant. We compared the WAIC of each model to a null-model without predictors (Intercept-model).

Model parameters were estimated using MCMC sampling, with 10000 iterations and 10000 tuning steps. The Bayesian regression yielded posterior distributions for each parameter of each participant. To evaluate significant effects, we considered 95% highest density intervals (HDI) of the *β* coefficient posterior distributions. We identified effects as significant when the 95% HDI indicated values either higher or lower than zero (95% HDI > 0 or 95% HDI < 0).

## 3. Results

### Random Dot Motion Task (RDM)

#### Convergence

We confirmed that all HDDM models converged, using visual inspection of MCMC trace plots and the Gelman-Rubin diagnostic whose values ranged between 0.9 and 1.1.

#### Model selection

In *Table 3*, we present the DICs of the HDDM models for RDM and their differences in DIC, compared to the *Null-model* (parameters not varing between groups/conditons) and the *HDDM-C* (parameters varying for motion coherence only). We performed two parallel model selections: one selection comparing the fits of these two control models with the fits of models including PDI as a predictor; and the other selection comparing the fits of the control models with those of models including CAPS as a predictor.

**Table 3.** DIC HDDMs. Deviance Information Criterion (DIC) for each HDDM in descending order; differences in DIC (Δ DIC) between each model and the Null-model (parameters do not vary for any condition/groups); Δ DIC between each model and the C-model (parameters vary only for motion coherence condition).

| | DIC | $\Delta$ DIC Null-model | $\Delta$ DIC HDDM-C |
| --- | --- | --- | --- |
| HDDM-TMS-CAPS-C | 9591.82 | -633.46* | -395.11* |
| HDDM-TMS-PDI-C | 9609.01 | -614.27* | -379.92* |
| HDDM-TMS | 9972.68 | -250.6* | -15.25* |
| HDDM-C | 9988.93 | -234.35* | 0.0 |
| Null-model | 10223.28 | 0.0 | 234.35 |
| HDDM-TMS-CAPS | 11216.66 | 992.38 | 1227.73 |
| HDDM-TMS-PDI | 11221.46 | 992.18 | 1232.53 |
| HDDM-TMS-C | 11857.4 | 1634.12 | 1868.47 |

\*DIC significantly lower

For the delusional phenotype, the model where parameters varied for the TMS, motion coherence, and PDI groups (i.e., *HDDM-TMS-PDI-C*) was the best model, outperforming the *Null-model*, the *HDDM-C* and the *HDDM-TMS* (parameters varying for TMS session). Similarly, *HDDM-TMS-CAPS-C* (parameter varying for TMS, motion coherence and CAPS groups) was the best model for the hallucinatory phenotype, outperforming the *Null-model*, *HDDM-C* and *HDDM-TMS*. Data simulation and parameter recovery for *HDDM-TMS-PDI-C* and *HDDM-TMS-CAPS-C* can be found in *Supplementary Materials*.

#### Group comparison

In *Fig 1,* we show distributions of posterior probability estimates of DDM parameters compared across motion coherence conditions, TMS sessions and psychotic phenotype groups in *HDDM-TMS-PDI-C* and *HDDM-TMS-CAPS-C*.

**Fig 1.**
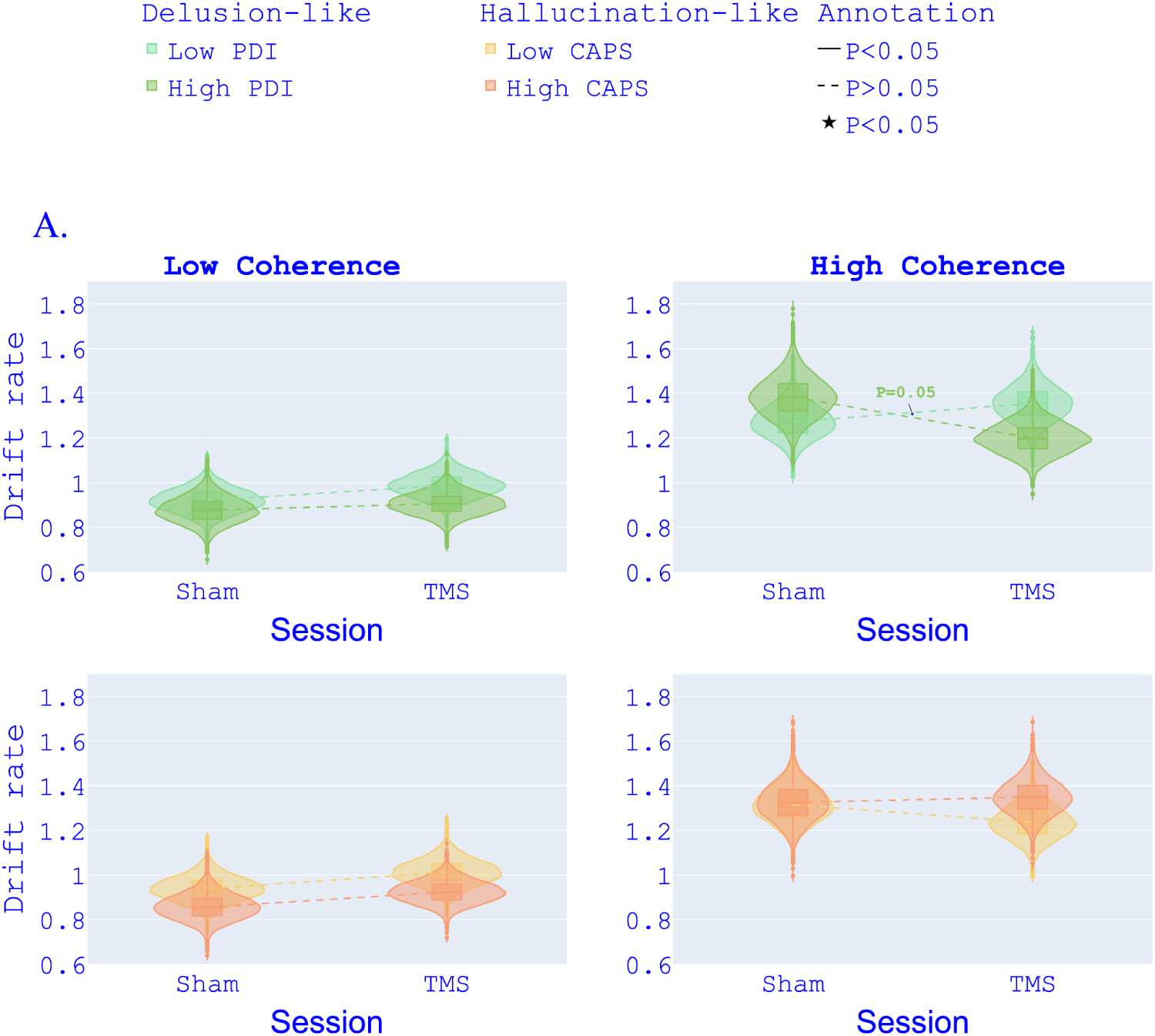
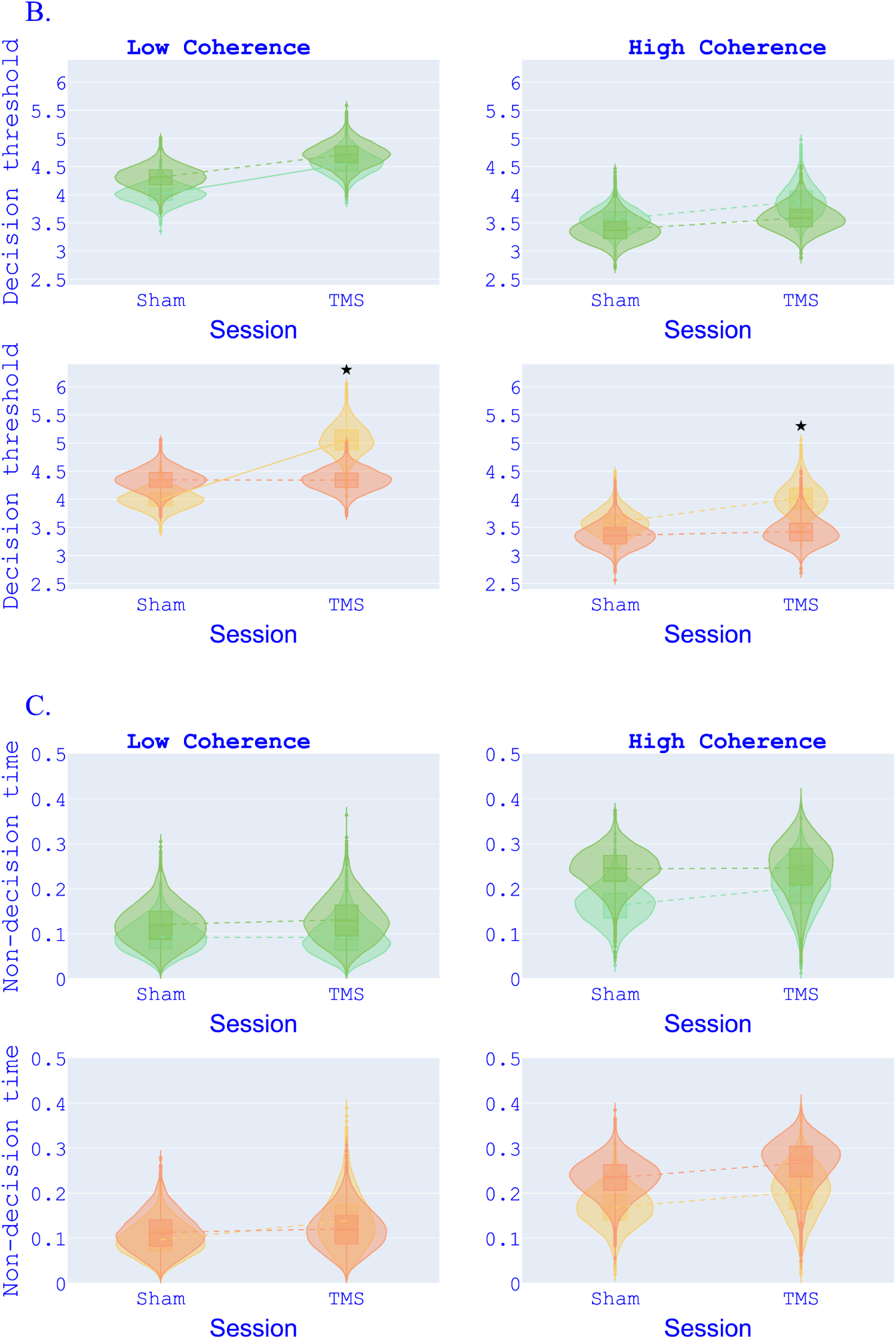
HDDM parameters for psychotic phenotype groups. Drift rate (A), decision threshold (B) and non-decision time (C) posterior probability distributions from HDDM-TMS-PDI-C (top) and HDDM-TMS-CAPS-C (bottom). Parameters are compared for different TMS conditions (Sham vs. TMS; unbroken lines) and for low and high motion coherence, PDI groups and CAPS groups (dashed lines).

The high-PDI group is the only group where drift rates appeared lower in TMS than in Sham, although the effect only lies on the threshold of significance (P=0.05) (see *Fig 1-A* top-right).

Decision threshold showed higher estimates in TMS compared to sham for low-PDI and low-CAPS groups in the low motion coherence condition (see Fig 1-B left). Decision threshold estimates for TMS were significantly different between low-and high-CAPS groups with low-CAPS showing higher decision thresholds (see Fig 1-B bottom). *HDDM-TMS-PDI-C* and *HDDM-TMS-CAPS-C* did not show any significant difference for non-decision time.

Data simulation and parameter recovery support the validity of these effects which were all recovered in synthetic models run with data simulated from *HDDM-TMS-PDI-C* and *HDDM-TMS-CAPS-C* parameter estimates (see *Supplementary materials*).

#### Post hoc HDDM-regression analysis

Given the results from the between-group comparison, we devised two HDDM models (*LR-HDDM-PDI* and *LR-HDDM-CAPS*) where drift rate, decision threshold and non-decision time varied as response variables of a linear regression. Model selection showed that the DIC of the *LR-HDDM-PDI* (DIC=11136.73) and the *LR-HDDM-CAPS* (DIC=11661.12) were higher (more poorly fitting) than the DIC of the *Null-model* (DIC=10223.28). The poor fit of the regression models was further confirmed by unsatisfactory predictive check and parameter recovery (see *Supplementary materials* for full report).

### Simple Reaction Time Task (SRT)

#### Mixed effect model

Our mixed effect model showed no evidence of TMS (M= 0.01; Q[0.025]= −0.05, Q[0.975]= 0.07; *p*=0.74), stimulus presentation (left or right) (M= −0.01; Q[0.025]= −0.05, Q[0.975]= 0.03; *p*=0.61) or their interaction (M= 0.02; Q[0.025]= −0.04, Q[0.975]= 0.08; *p*=0.47) on RTs in SRT.

#### HDDM modelling

We ran post hoc HDDMs, namely *HDDM-SRT-Null* and *HDDM-SRT-TMS* to further investigate TMS effects on DDM parameters. The DIC of *HDDM-SRT-TMS* (DIC= −2786.31) was not significantly higher than the DIC of the *HDDM-SRT-Null* (DIC= −2780.78), showing that TMS does not add explanatory value to HDDM modelling of SRT data.

### Beads Task (BT)

From the linear mixed model analysis, we did not find any significant effect of TMS manipulation on DTD in the BT (M= −0.99; Q[0.025]= −2.05, Q[0.975]= 0.08; *p*=0.07). This non-significant effect of TMS was confirmed by the hierarchical Bayesian Poisson regression. All linear models with predictors outperformed the *Intercept-model* (see *Table 4*).

**Table 4.** WAIC Beads task regression models. Watanabe–Akaike information criterion (WAIC) for each regression models predicting draws-to-decision in the Beads task; differences in WAIC (Δ WAIC) between each model and the Null-model (where parameters do not vary for any condition/groups)..

| | WAIC | $\Delta$ WAIC Intercept-model |
| --- | --- | --- |
| **LM-TMS** | -2759.2 | **-110.48\*** |
| **LM-TMSxCAPS** | -2759.1701 | **-110.8001\*** |
| **LM-TMSxPDI** | -2759.1744 | **-110.8048\*** |
| **Intercept-model** | -2648.37 | 0.0 |

\*WAIC significantly lower

However, we did not find any effect of TMS (M= −0.94; Q[−1.95]= −1.96, Q[0.97]= 0.11), nor of PDI (M= −0.001; Q[0.03]= −0.002, Q[0.97]= 0.01), CAPS (M= −0.001; Q[0.03]= −0.02, Q[0.97]= 0.01), or any TMS interaction with PDI (M= 0.001; Q[0.03]= −0.02, Q[0.97]= 0.03) or CAPS (M= 0.004; Q[0.03]= −0.02, Q[0.97]= 0.03) on DTD. From visual inspection of MCMC trace plots and built-in diagnostics, both the GLMM and the hierarchical Bayesian regression showed good convergence.

### Between-group demographics and psychotic phenotype

*Table 5* shows differences in demographics, PDI and CAPS scores between TMS conditions, PDI and CAPS median-split groups.

**Table 5.**
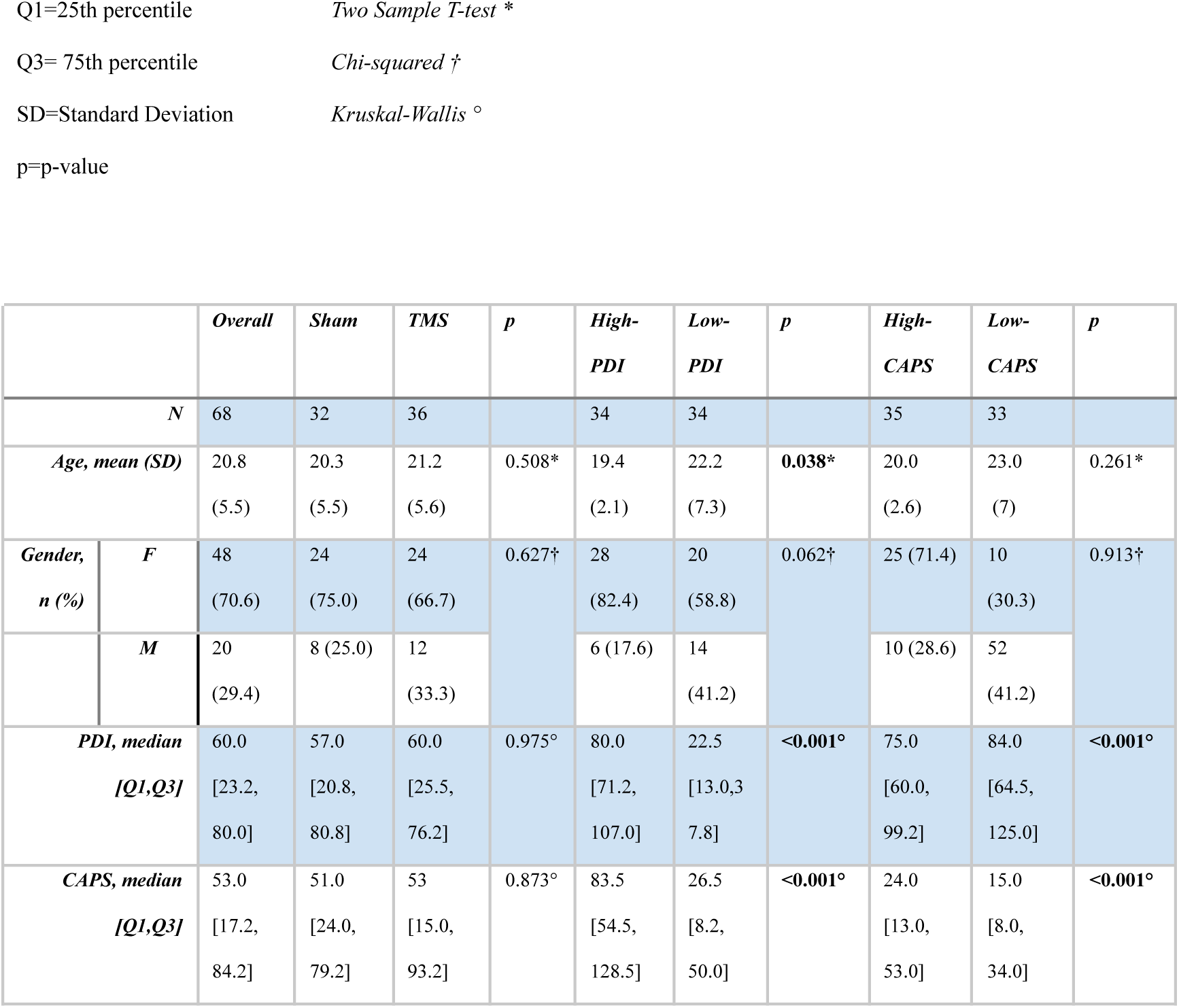
Demographics and psychotic phenotype compared between TMS sessions and psychotic phenotype groups.

There was no significant difference in age, gender, PDI or CAPS scores between sham and TMS conditions. We found significantly higher PDI scores in the high-CAPS group compared to the low-CAPS group and, correspondingly, CAPS scores were higher in the high-PDI group compared to the low-PDI group. Finally, we found age was significantly higher in the low-PDI group compared to the high-PDI group.

## 4. Discussion

We applied offline 1 Hz TMS over right PPC and compared group-level estimates of DDM parameters against a control group that received a sham TMS session. We evaluated the effect of TMS across groups of participants obtained by scores median-split by delusional phenotype (high- vs. low-PDI) and hallucinatory phenotype (high- vs. low-CAPS).

The models that best fitted RDM data showed increased decision thresholds in the TMS session compared to sham in participants with low psychotic phenotypes (low-PDI and low-CAPS groups). Remarkably, participants with higher hallucinatory phenotypes showed instead stable values of this parameter across TMS sessions. Disruption of right PPC activity notably diminished the precision of sensory evidence (as proxied by drift rate) solely in participants exhibiting a high delusional phenotype, with an effect reaching the edge of significance.

Importantly, the TMS manipulation did not affect the performance on a control decision task (SRT) where no accumulation of ambiguous evidence was involved. Nor did we find any effect of TMS on DDM estimates of the non-decision time parameter which approximates the time in the RT taken by non-decisional processes (i.e., stimulus encoding and motor execution).

### Disruption of right PPC in low psychotic phenotype decreases decision thresholds

Evidence from neurophysiological studies shows that neuronal activity in IPS in humans and LIP in non-human primates correlate with the tally of accumulated evidence but not with the discrimination of coherent motion involved in RDM^20,41,42^. In line with this literature, our results on the low psychotic phenotype show that reducing the excitability of the cortical area around the right PPC (in the vicinity of IPS) affects the tally of accumulated evidence (i.e., decision threshold) but not the precision encoding of coherent motion information (i.e., drift-rate).

By decreasing cortical excitability, TMS may have prompted PPC neurons to require more excitatory input from the visual cortex to initiate behaviour. This aligns with findings suggesting PPC neurons accumulate sensory evidence until reaching a firing rate threshold before making a decision^20,42–44^. TMS may have reduced the baseline excitability of PPC, requiring more excitatory signalling to reach the stereotypical firing rate, thus increasing decision thresholds. Concurrently, evidence suggests sensory information accumulation in PPC may be regulated by oscillatory activity, acting as a filter in a process described as the “parietal bottleneck”^20,45,46^. 1 Hz TMS may disrupt this activity, potentially increasing decision thresholds. To further test this latter interpretation, research on the “parietal bottleneck” hypothesis could explore PPC TMS effects on oscillatory activity and DDM parameters.

### No effect of TMS on decision threshold in high psychotic phenotype

Given the account above, if TMS reduced decision thresholds by down-regulating baseline activity in people with low hallucinatory phenotype, the lack of this effect in the high-CAPS group might indicate a higher baseline level of PPC excitability in that group, allowing neurons to reach their canonical firing rate threshold despite the TMS. This account may find support in the evidence showing hyperactivity of sensory cortices in hallucinating patients^47,48^ and in the population experiencing anomalous perceptions^49^. However, no evidence has yet suggested that this is the case for associative areas like PPC. Our findings raise the question of whether the hallucinatory phenotype is also associated with increased cortical activity in areas involved in decision-making such as the PPC.

This finding acquires aetiological interest when considering the evidence linking PPC activity and the emergence of perceptual conscious experiences. Reaching a critical threshold of firing rate in PPC neurons might underlie the perceptual experience of a stimulus. Some compelling evidence for this comes from Pereira et al. (2020) who recorded single-neuron activity in the PPC of an epileptic patient performing a tactile signal detection task^50^. Neurons in PPC showed the typical ramping activity for tactile stimulations that were detected while firing rates were lower for those stimuli that did not trigger a conscious experience. Supporting this correlational evidence, TMS pulses over the IPS induce perceptual fading of stimuli presented peripherally in the visual field^51^. A higher baseline of PPC excitability might predispose an individual to experience percepts with a lower amount of excitatory signal coming from sensory areas. In this way, neural activity non-induced by external stimuli would be more likely to contribute to the emergence of anomalous conscious percepts.

### TMS over right PPC reduces sensory precision in the delusional phenotype

We showed that TMS over the right PPC decreased sensory precision (proxied by drift rate) only in individuals with a higher delusional phenotype. This result can be understood in light of PCA and dysconnectivity associated with psychosis and proneness to psychosis^52,53^. When performing probabilistic decision tasks where the accumulation of evidence is required, patients with schizophrenia show abnormal recruitment of regions within the task-positive network (including PPC) and the default mode networks^54–56^. The disruption of PPC neuronal activity by TMS in people with a higher delusional phenotype might have exacerbated aberrant connectivity patterns in the network that could be already present in people prone to delusion. The specificities of these dysconnectivity patterns are a matter for further investigation.

### PPC role in perceptual vs. cognitive inference

Unlike RDM, BT performance was not affected by PPC disruption differently between individuals with low and high psychotic phenotypes. These results support the idea that mechanisms of perceptual rather than cognitive inference might underlie key differences in neurocomputational mechanisms along the psychotic phenotype.

Although some evidence from correlational studies associated IPS activity with the commitment to choice in probabilistic decision tasks^32,57^, we did not find any significant effect of TMS on the quantity of collected evidence in BT. We cannot exclude that with a larger sample, offline TMS on PPC could affect data-gathering in tasks requiring higher cognitive functions.

Psychotic phenotype scores also did not affect BT performance in line with previous studies on the general population^29,58–60^. Specifically in our previous work^29^, we already suggested that the BT might not be the ideal paradigm for testing psychotic phenotype impact on evidence accumulation. A primary concern with the task is the number of trials typically used in the BT where often designs include only one trial^61–63^. This is because the jumping-to-conclusions bias associated with delusional symptomatology appears to decay after a few trials^62^. However, a limited number of trials per subject makes measurements more susceptible to random variance in performance. We addressed this concern using 20 trials per subject and we did not find any data-gathering bias associated with the psychotic phenotype. On the other hand, the RDM performance can be evaluated over several trials per subject, offering a higher test-retest reliability^64^.

## Limitations and conclusions

One key limitation of our study involves the method we used to target IPS. Although P4 in the 10-20 system can reliably target the cortical region around IPS^36^, using neuronavigation through individual MRI scans can offer a more precise anatomical localisation. A second matter to be aware of when considering our findings is that offline TMS did not disrupt cortical activity at precise time points but had an effect along the whole task execution. The 1 Hz single pulses coupled with stimulus encoding might or might not yield the effect we found on RDM. Finally, we explored the effects only of unilateral TMS on the rIPS of right-handed participants. Future studies could expand our findings by testing whether bilateral IPS inhibition affects: drift rates irrespective of delusional phenotype, decision threshold in participants with higher hallucinatory phenotype, or DTD in BT.

Altogether, our findings suggest that PPC plays a pivotal role in terminating evidence accumulation in perceptual decisions and that this area might be involved in decision-making and computational patterns associated with psychosis. Neural mechanisms localised in PPC or that involve this area on a network level might be of relevance for changes in inferential processes in the psychotic phenotype. Our results showed that the disruption of the right PPC affects delusional and hallucinatory phenotypes differently, suggesting that the mechanisms involving PPC might be different for the two psychotic phenotypes.

## Data Availability

All data produced are available online at:
https://github.com/frscaramozzino/Posterior-Parietal-Cortex-Modulates-Perceptual-Decisions-Depending-On-Psychotic-Phenotype

https://github.com/frscaramozzino/Posterior-Parietal-Cortex-Modulates-Perceptual-Decisions-Depending-On-Psychotic-Phenotype

## Acknowledgements and disclosures

This work was supported by a grant from the *NOMIS Foundation* (“Collective Delusions: Social Identity and Scientific Misbeliefs”). The authors declare that they have no financial or personal relationships that could influence the work reported in this article.

## Supplementary materials

### Distributions of psychotic phenotype measures

To provide a comprehensive representation of the data on the psychotic phenotype within our sample, we present the medians and frequency distributions as a percentage of the sample for both PDI and CAPS in *Figure 1-A-B*.

**Fig 1.**
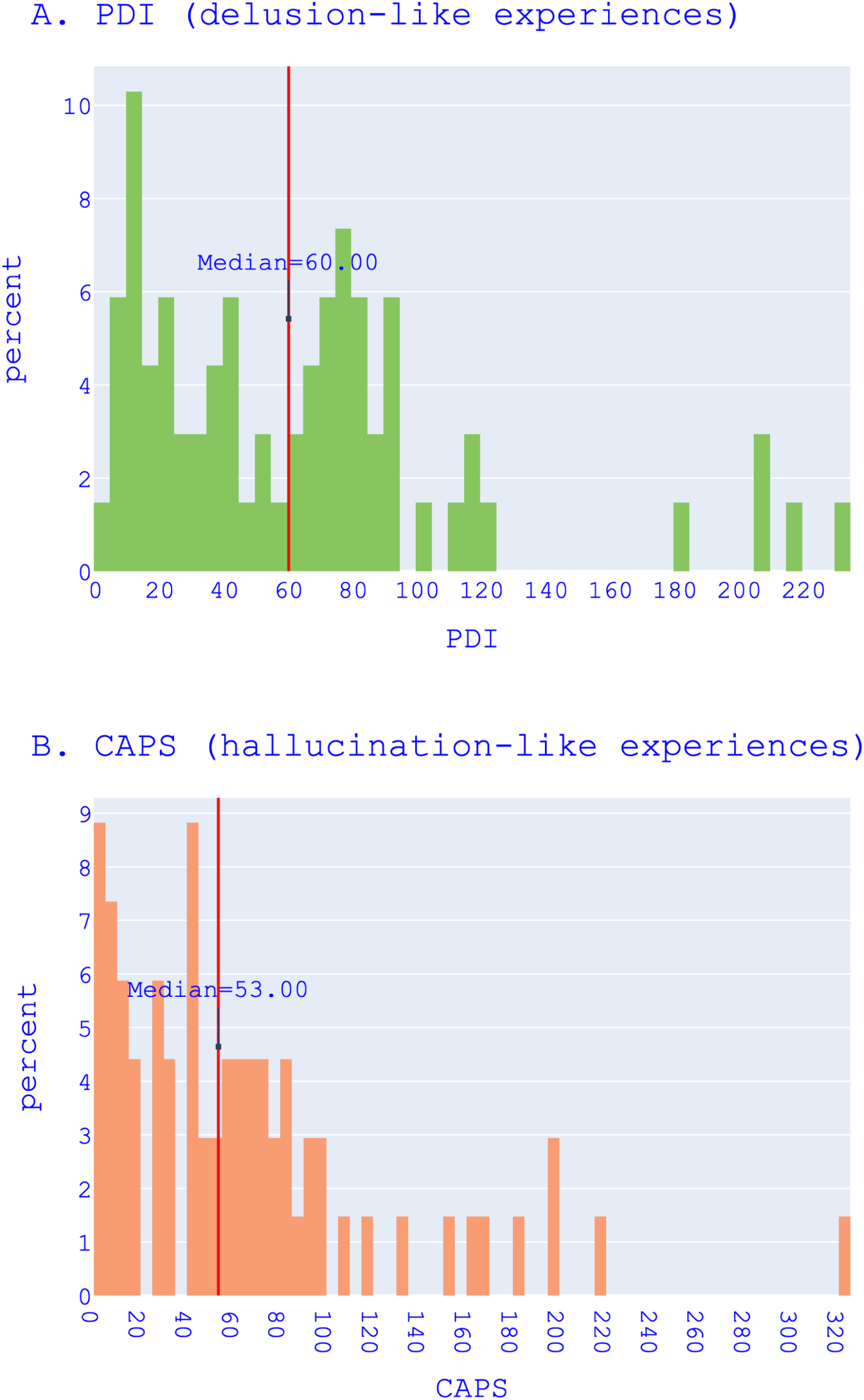
Frequency distribution of psychotic phenotype measures. Distribution in participant percentage of (A) Peters et al. Delusion Inventory (PDI), (B) Cardiff Anomalous Perceptions Scale (CAPS).

### Data Simulation and Parameter Recovery

We provide here data simulation and parameter recovery of the winning hierarchical drift-diffusion models (HDDMs) fitted to the data from the Random Dot Motion task (RDM). We performed data simulation and parameter recovery of the winning between-group models *HDDM-TMS-PDI-C* and *HDDM-TMS-CAPS-C* and of regression models *HDDM-LR-TMS-PDI* and *HDDM-LR-TMS-CAPS*. We simulated the data using the *hddm.generate.gen_rand_data(…)* function of the *HDDM 0.8.0* package (Wiecki et al., 2013). In *Figure 2*, we show the distribution of RTs inputted to these models.

**Fig. 2.**
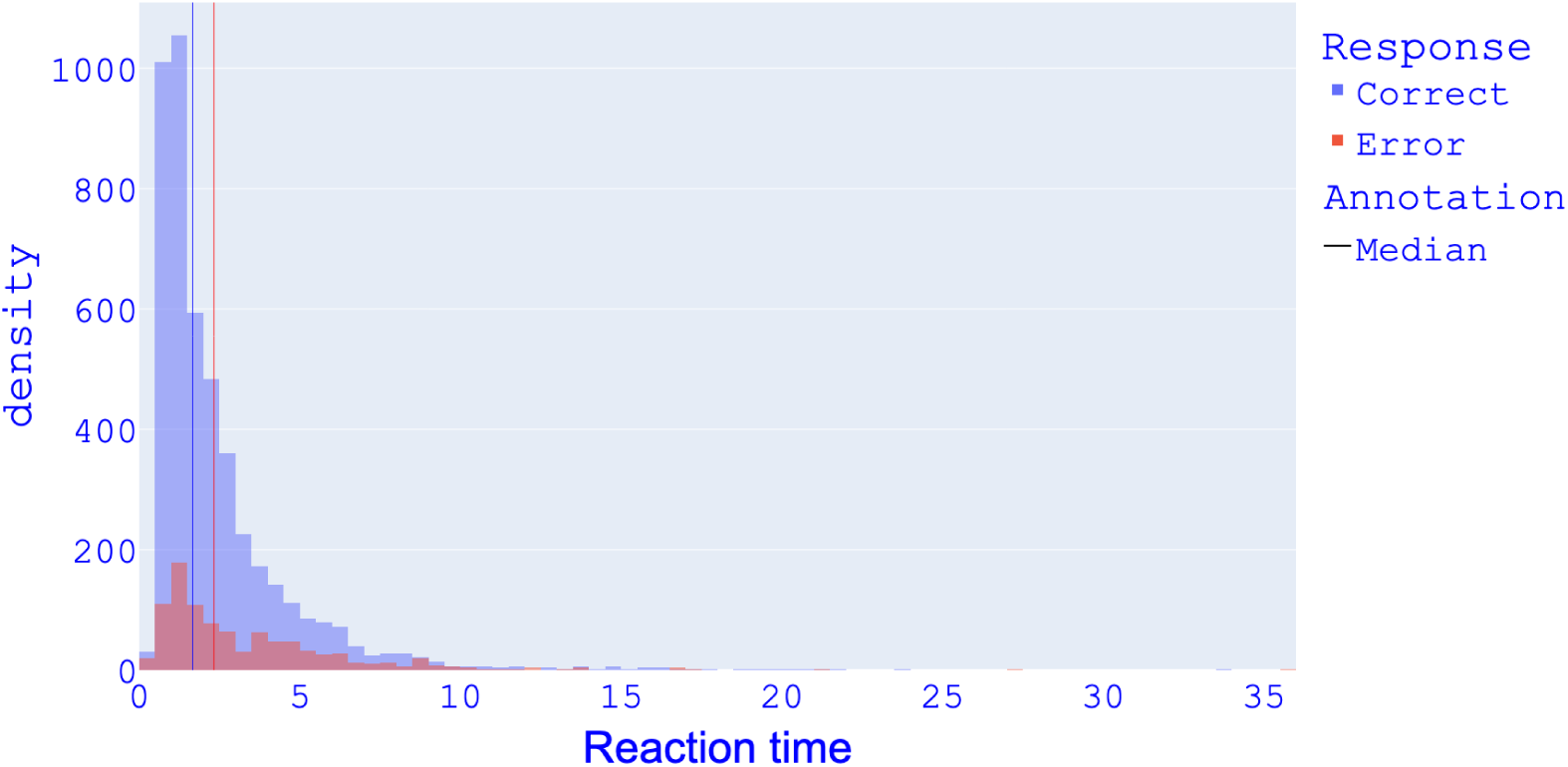
RT distribution. Distribution of reaction times (in seconds) for correct and error responses as inputted to HDDMs; fast outliers cut-off: 0.2 s; no slow outliers cut-off.

#### Between-group HDDMS

HDDM-TMS-PDI-C and HDDM-TMS-CAPS-C show a mean root-mean-square error (RMSE) of 0.283 s for correct responses. The RT predictions of the same models were less accurate for error responses with a mean RMSE of 2.621 s (see *Figure 3*).

**Fig. 3.**
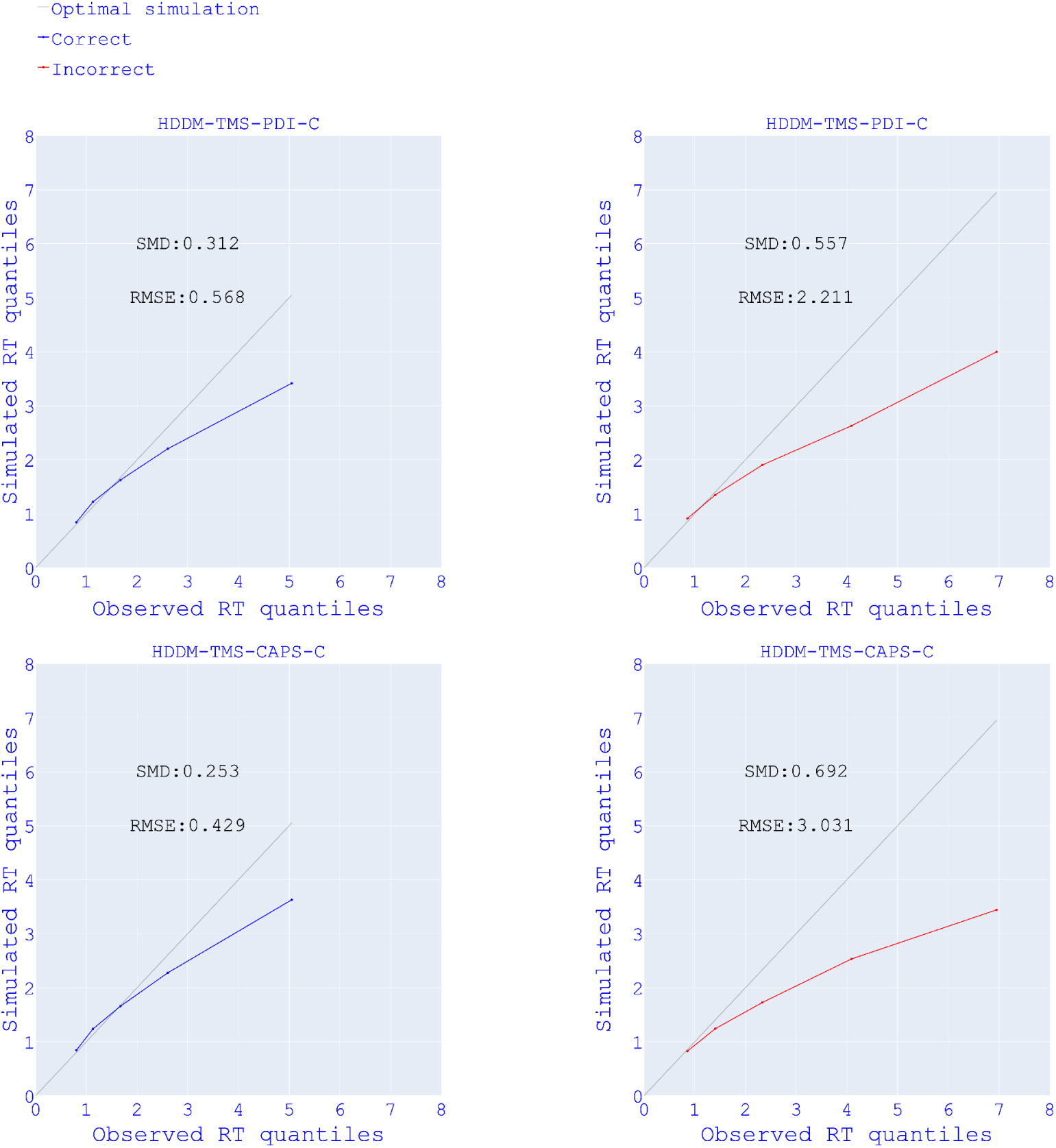
Reaction time (RT) predictions of between-group HDDMs. Linearity plots of observed RT quantiles (x-axis) and simulated RT quantiles (y-axis) for correct and error responses of between-group models in *Study 2*. SMD= Standardised mean difference; RMSE= Root mean squared error.

The between-group models’ recovered drift rates were slightly overestimated compared to the empirical models’ estimations (see *Figure 4* and *Figure 5*). This overestimation generally did not affect the recovery of the differences between the sham and TMS groups: the difference in drift rate present on the threshold of significance in the empirical model was recovered in the high-PDI group (significant in the simulation model; see *Figure 5*, top-right). The simulation models reproduced the effect of TMS on decision thresholds in low-PDI and low-CAPS groups in the low motion coherence (see respectively *Figure 4* and *Figure 5* Low Coherence centre-left). Non-decision times were generally well recovered.

**Fig. 4.**
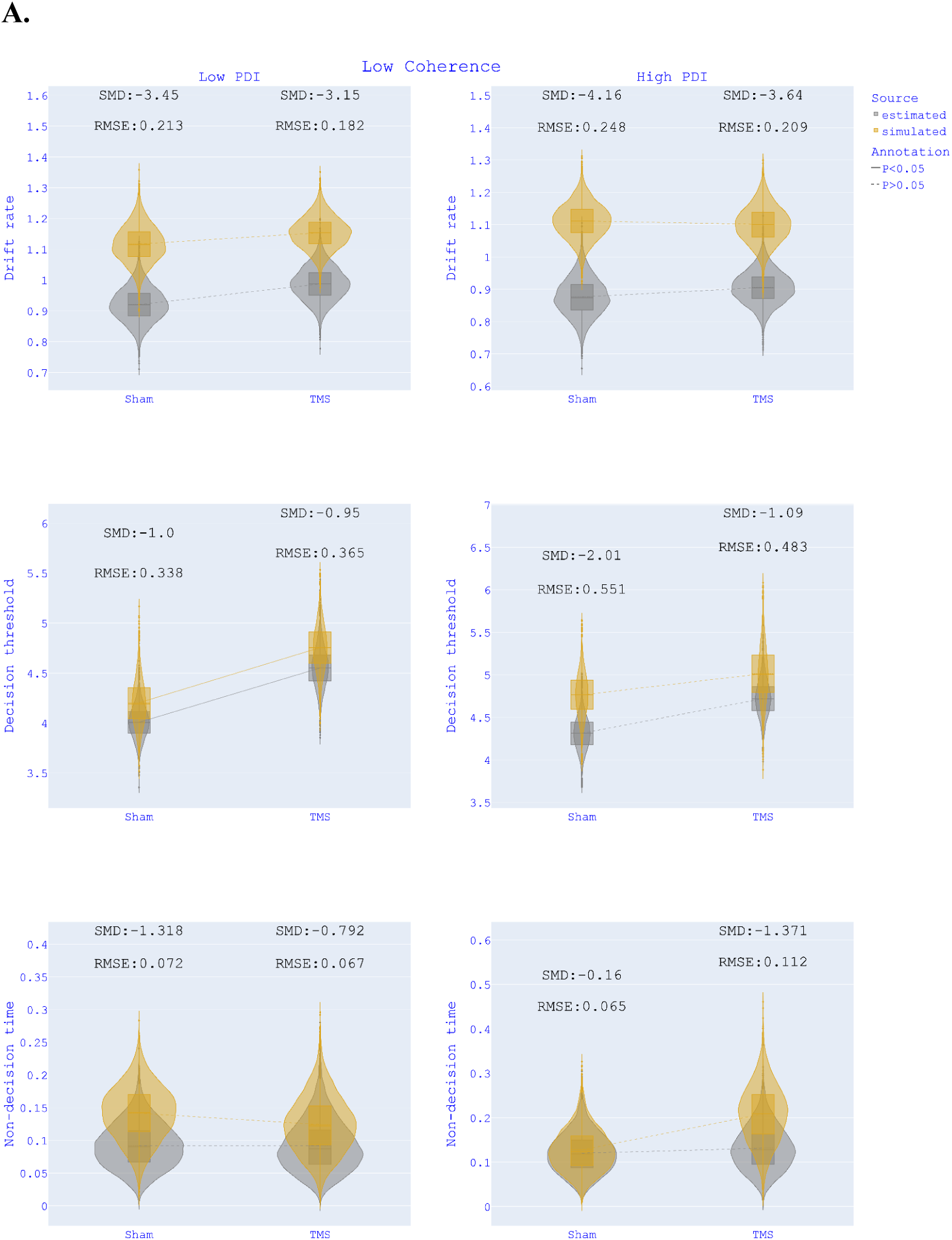
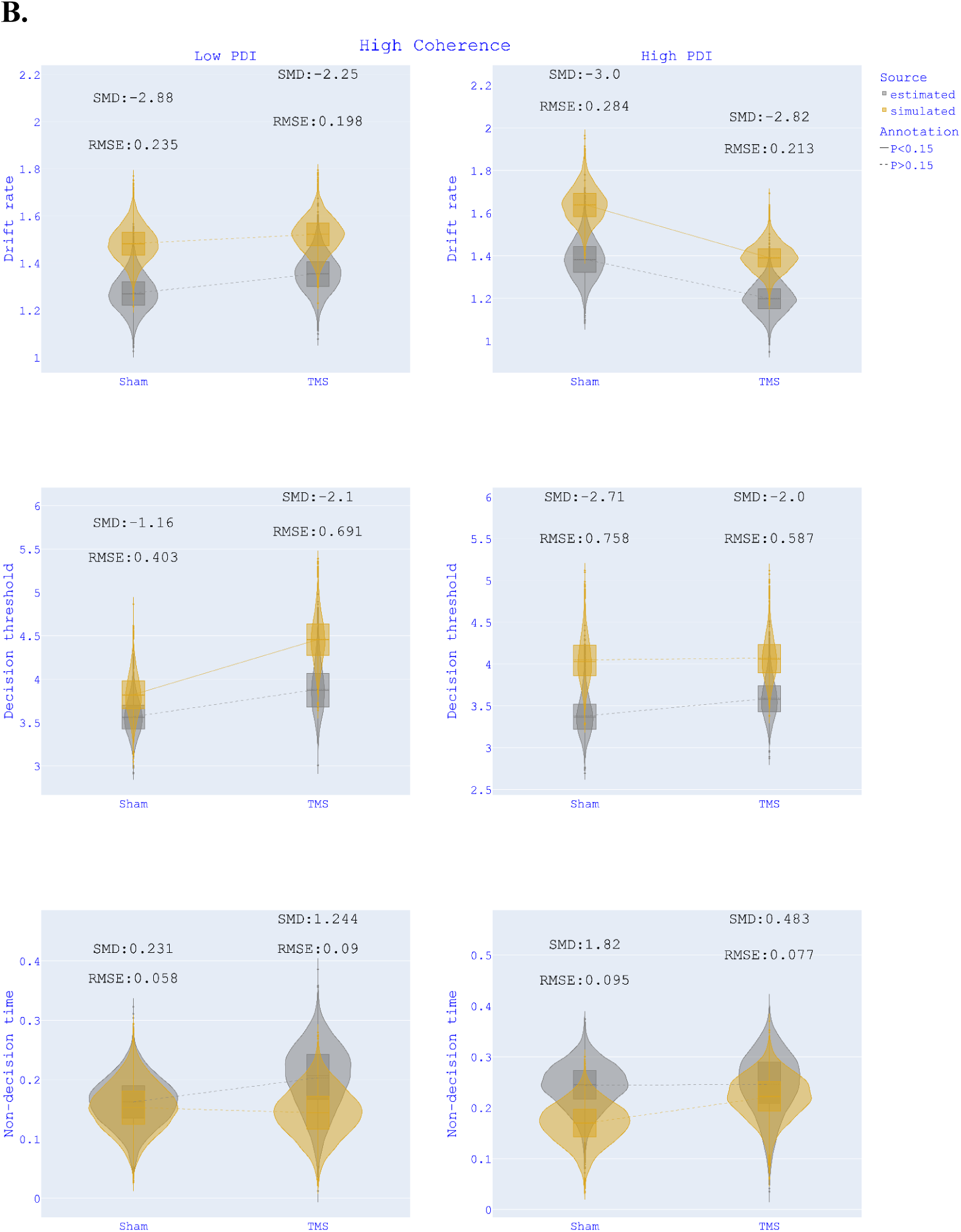
Parameter recovery HDDM-TMS-PDI-C. Recovery of drift rates, decision thresholds and non-decision times for HDDM-TMS-PDI-C (TMS session - PDI groups - Motion Coherence) divided by Low Coherence (A) and High Coherence (B) estimates. SMD = Standardised mean difference; RMSE = Root mean squared error.

**Fig. 5.**
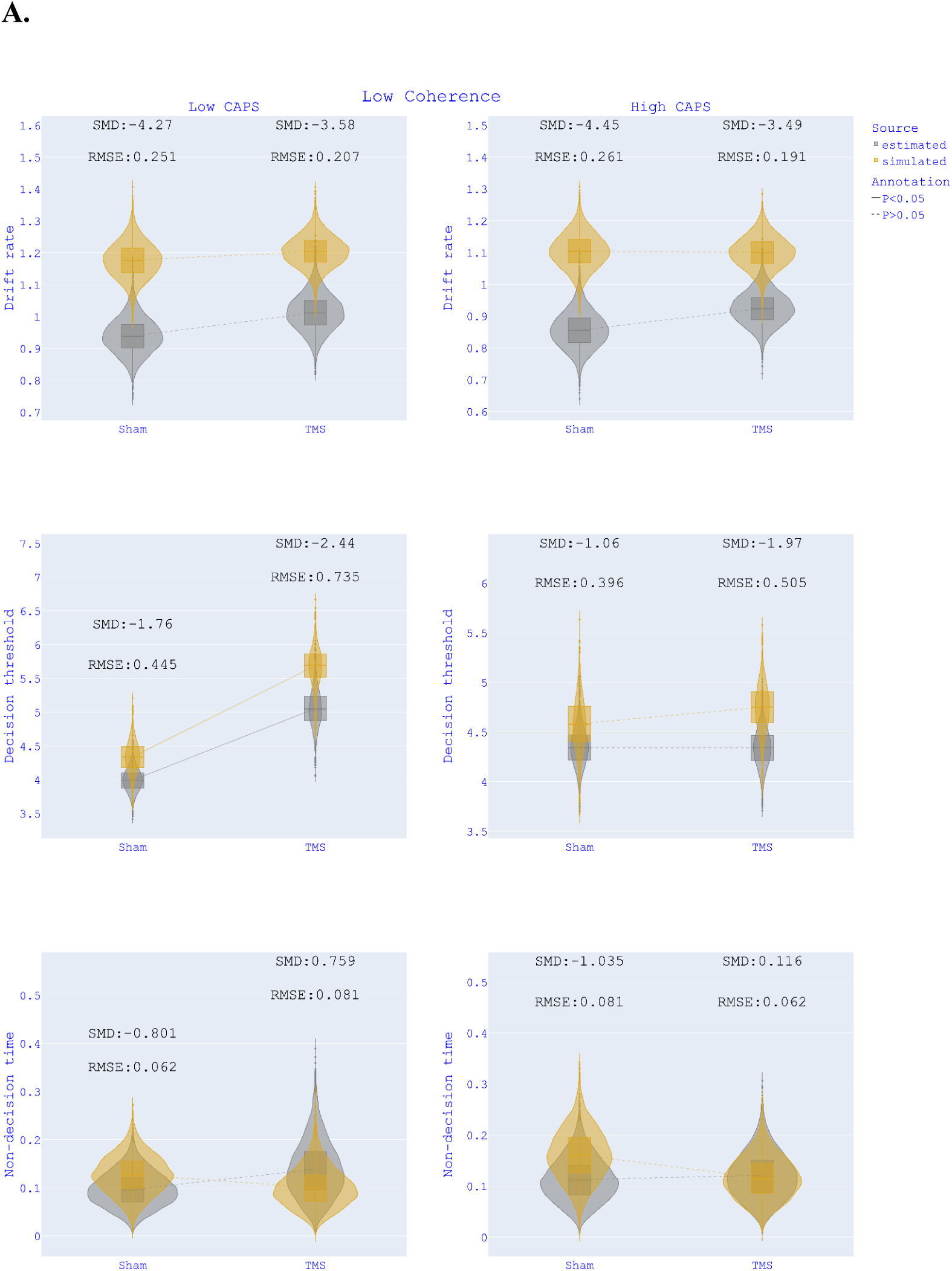
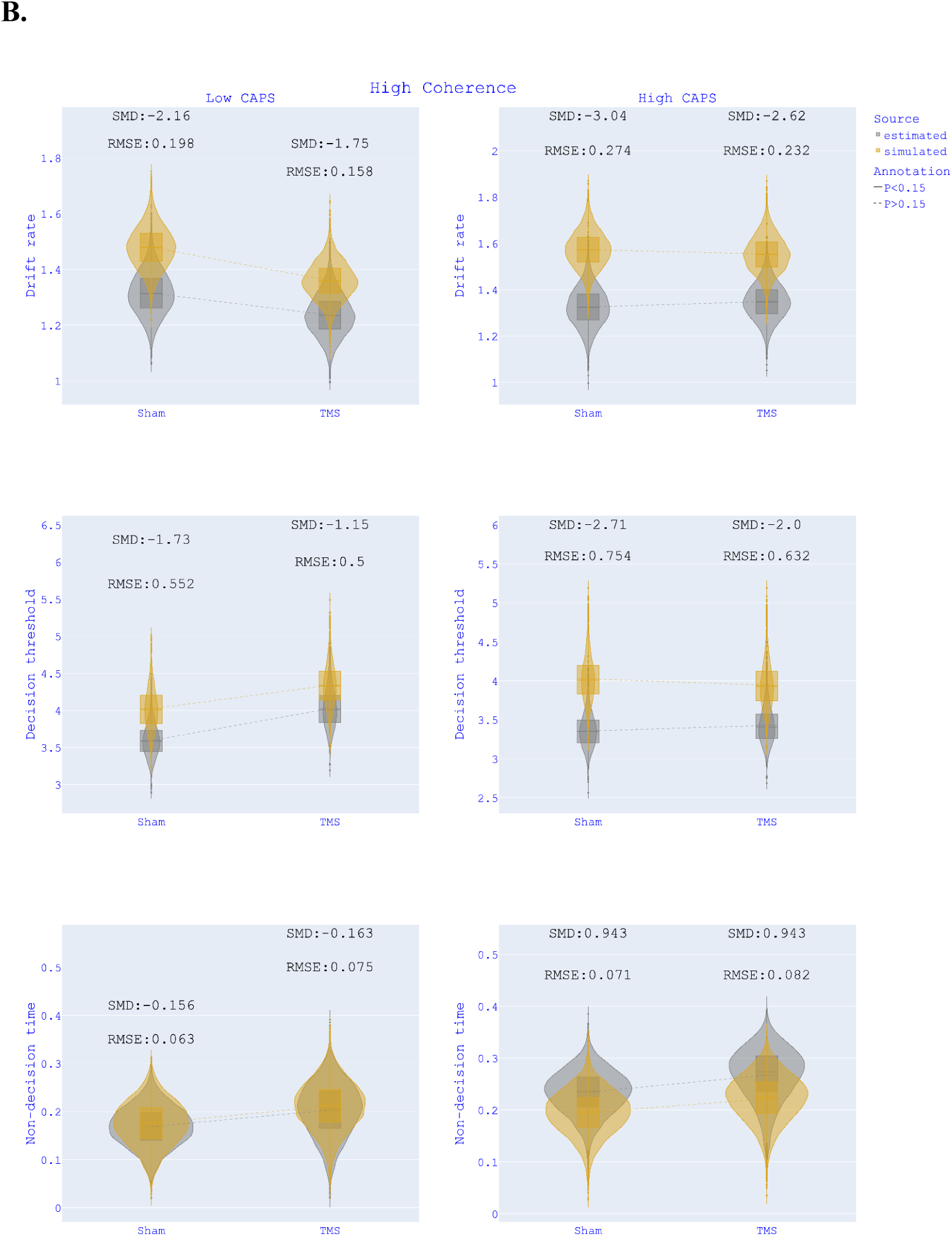
Parameter recovery HDDM-TMS-CAPS-C. Recovery of drift rates, decision thresholds and non-decision times for HDDM-TMS-CAPS-C (TMS session - CAPS groups - Motion Coherence) divided by Low Coherence (A) and High Coherence (B) estimates. SMD = Standardised mean difference; RMSE = Root mean squared error.

#### Regression HDDMs

Motion coherence, TMS, PDI and PDIxTMS were predictors of HDDM parameters in model *LR-HDDM-PDI*; motion coherence, TMS, CAPS and CAPSxTMS in *LR-HDDM-CAPS*. These HDDM regression models showed highly imprecise predictions of quantiles for both correct and error responses (all RMSEs over 2 s; see *Figure 6*). In *Figure 7*, we show the recovery of *β* coefficients for the interaction terms between the psychotic phenotypes and TMS session which tested one of our main hypotheses for regression HDDMs. The positive effect of the TMS-CAPS interaction was well-recovered, while the same effect for PDI was not reproduced by the simulation model. For non-decision times, the negative interactions of TMS with respectively PDI and CAPS were not reproduced by any simulation model. The empirical *β* coefficients were well-recovered for the decision threshold by both simulation models.

**Fig. 6.**
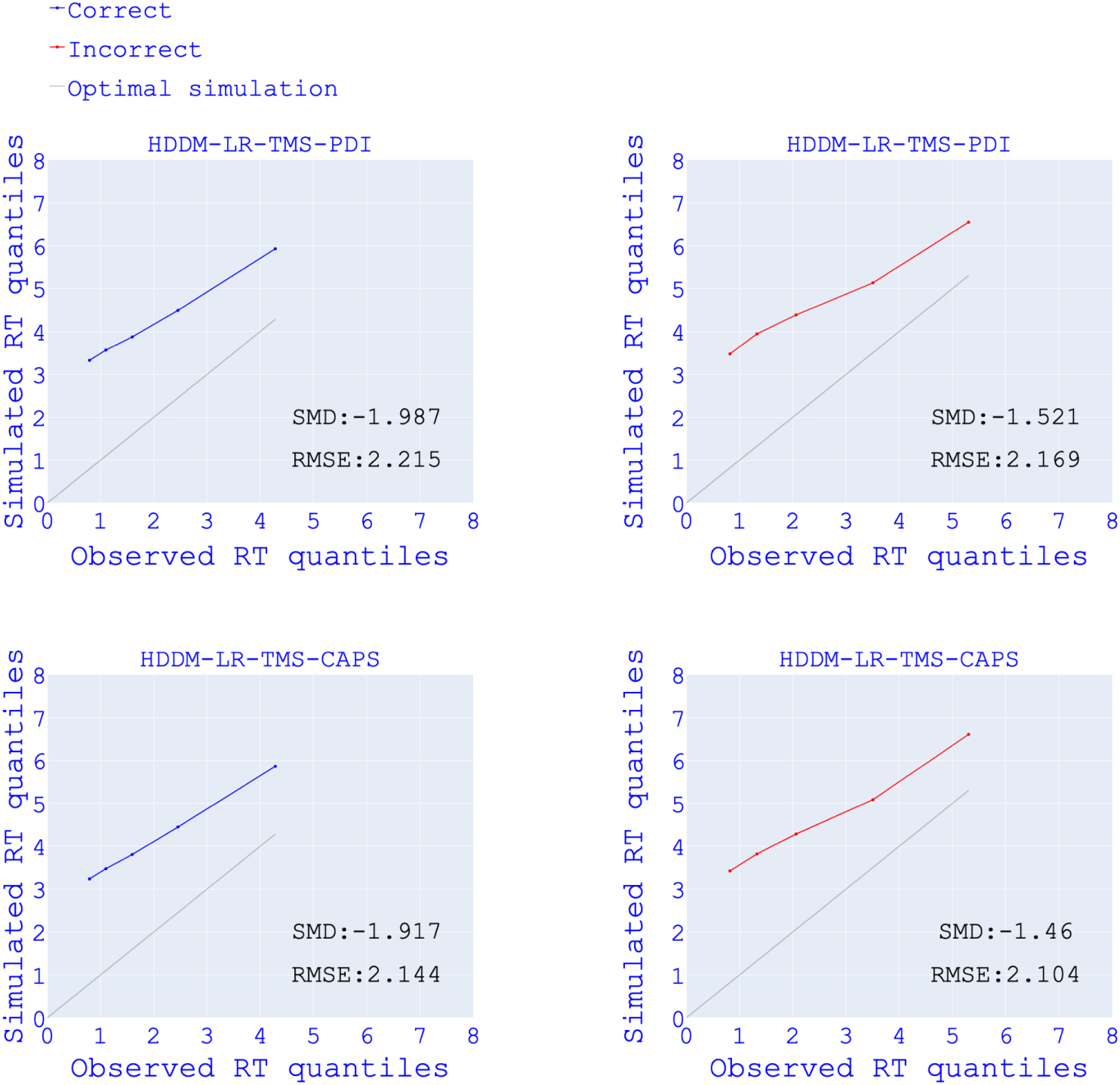
RT predictions of regression HDDMs. Linearity plots of observed reaction time (RT) quantiles (x-axis) and simulated RT quantiles (y-axis) for correct and error responses of regression models. SMD = Standardised mean difference; RMSE = Root mean squared error.

**Fig. 7.**
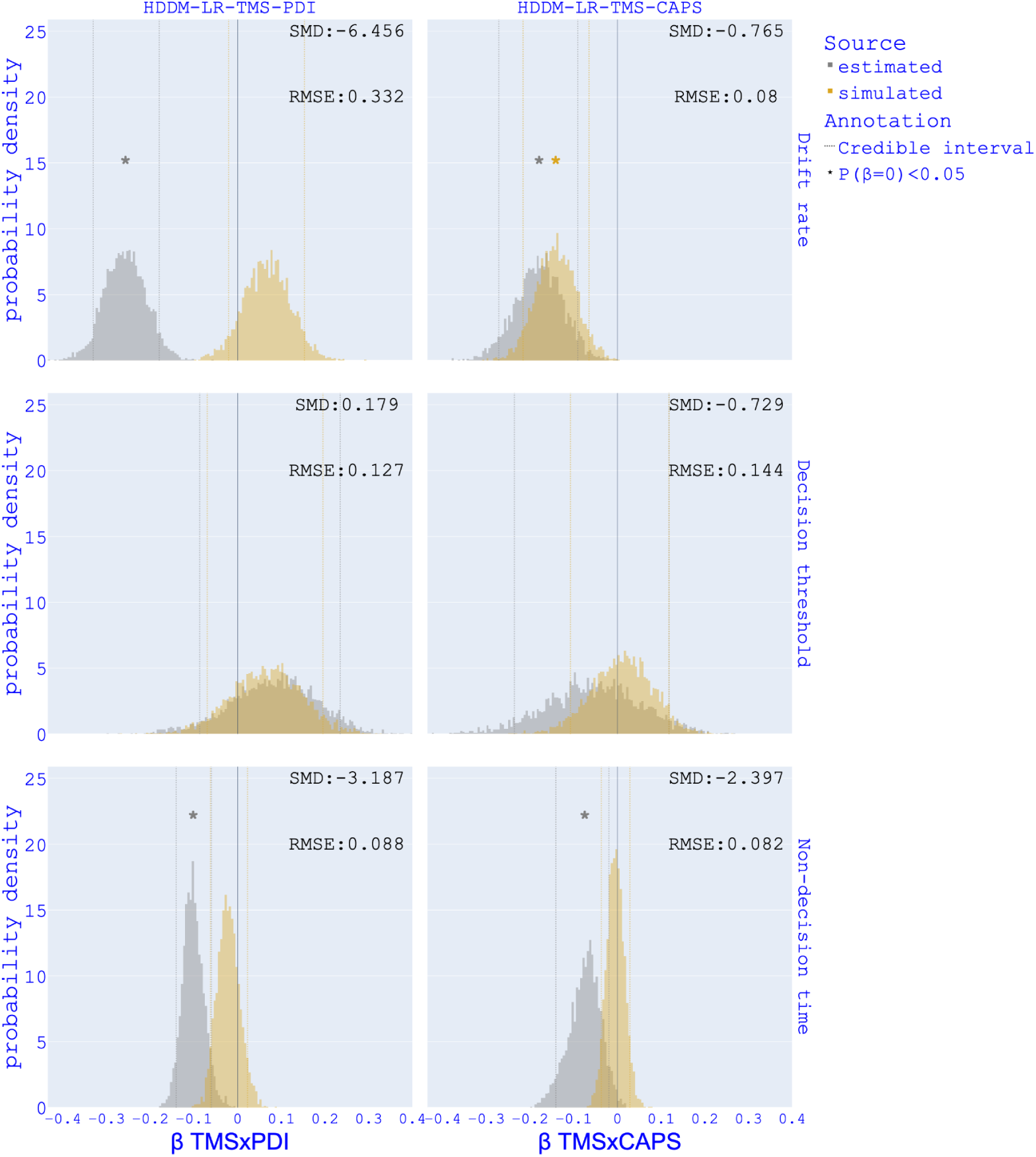
Parameter recovery for TMS interaction with psychotic phenotypes HDDM-LR -TMS-PDI and HDDM-LR-TMS-CAPS. Recovery of β for TMS x Schizotypy in HDDM-LR -TMS-PDI (TMS session - PDI - interaction TMSxPDI), HDDM-LR-TMS-CAPS (TMS session - CAPS - interaction TMSxCAPS). SMD= Standardised mean difference; RMSE= Rooted mean squared error.

